# Acute myeloid leukemia risk stratification in younger and older patients through transcriptomic machine learning models

**DOI:** 10.1101/2024.11.13.24317248

**Authors:** Raíssa Silva, Cédric Riedel, Maïlis Amico, Jerome Reboul, Benoit Guibert, Camelia Sennaoui, Chloé Bessiere, Florence Ruffle, Nicolas Gilbert, Anthony Boureux, Thérèse Commes

## Abstract

Acute Myeloid Leukemia (AML) is a genetically and clinically heterogeneous disease that can develop at any age. While AML incidence increases with age and distinct genetic alterations are observed in younger versus older patients, current classification systems do not incorporate age as a defining factor. In this study, we analyzed RNA-seq data from 404 AML patients at initial diagnosis, leveraging a k-mer-based machine learning approach to uncover age-related transcriptomic differences in favorable and adverse risk groups. Our model achieved over 90% accuracy in risk prediction and identified key gene signatures distinguishing ELN2017 favorable and adverse groups. From these signatures, we selected prognostic biomarkers with significant impacts on survival. Additionally, we explored the biological context underlying transcriptomic complexity across age groups, revealing distinct tumor profiles and differences in immune and stromal cell populations, particularly in older patients. These findings underscore the importance of age-related molecular features in AML and provide new insights for risk stratification and therapeutic targeting.

## Introduction

Acute Myeloid Leukemia (AML) is a heterogeneous and complex disease with variations in morphology, immunophenotype, genetic, epigenetic signatures, leading to different responses to treatment ^1^. Current classification systems, such as the European LeukemiaNet (ELN), stratify AML patients into favorable, intermediate, and adverse risk categories based on cytogenetic profiles and molecular biomarkers at the genomic DNA level.

A recent study ^2^ presents the revised 2022 ELN genetic-risk classification as suitable for prognostic stratification of patients with AML. However, it highlights the importance of distinguishing younger patients (≤ 60 years) from older patients (> 60 years), given the differences in genetic alterations and clinical outcomes.

Although AML can occur at any age, studies have described age as an important factor in the prognosis of AML ^3^, and its management presents more challenges as age increases ^4^. Eisfeld et al.^5^ proposed incorporating genetic profiling to improve the stratification of older patients (>60 years), a finding later supported by Mims et^6^ and Li et al.^7^. However these studies did not explore gene expression profiles to improve the risk classification in younger and older patients separately. Considering the need to improve the AML classification, this study aims to investigate differences between the age groups in ELN2017 classification by analyzing the RNA-sequencing (RNA-seq) data.

RNA-seq is an accurate method for transcriptome profiling, allowing the identification of key molecular signatures that can drive disease pathogenesis and progression. As proposed by Docking et al. ^3^, RNA-seq has the potential to serve as a standalone assay for both AML diagnosis and prognosis, offering a deeper understanding of patient subgroups based on their transcriptomic profiles.

We recently demonstrated that RNA-seq data can be analyzed using k-mer based approaches, which, unlike traditional methods guided by reference annotations, are reference-free and do not require pre-mapping or gene counting ^8,9^. This approach fragments sequencing reads into k-mers, substrings of length k, that are indexed to provide a compressed yet comprehensive representation of the data. This method enables efficient exact and approximate sequence searches ^10,11^ while significantly reducing computational time ^12^. The k-mers approach offers rapid and large-scale analysis and can be powerful for applying machine learning models.

Integrating Machine Learning (ML) and k-mer-based approaches presents a promising strategy for capturing the complex biological landscape of transcriptomes and refining patient risk classification. ML methods offer new opportunities in this field, the integration of ML and RNA-seq has recently shown its effectiveness for prognosis in cancer, mainly based on counted gene expression.

In this study, we applied ML models trained on k-mer count matrices to predict favorable and adverse risk groups in AML at initial diagnosis for younger and older patients. Next, we analyze the difference between favorable and adverse risk in the two age groups and we provide a list of genes of interest for AML and the impact on survival. We also analyzed biological features that distinguish the older patients in relation to risk prediction.

## Materials and methods

### Transcriptome data and clinical information

We analyzed six transcriptome cohorts of AML patients. To investigate ELN2017 risk stratification, we analyzed 404 samples with favorable or adverse risk: 212 samples from Beat-AML ^13^; 112 samples from Beat-AML2.0 ^14^; and 80 samples from Leucegene ^15^. We also performed survival analysis using 240 samples from the Beat-AML and 37 from the GSE62852 cohort for training survival models, and 129 samples from the EGAD00001006701 cohort for testing. Table 1 presents an overview of risk stratification and age groups from the AML cohorts.

**Table 1.**
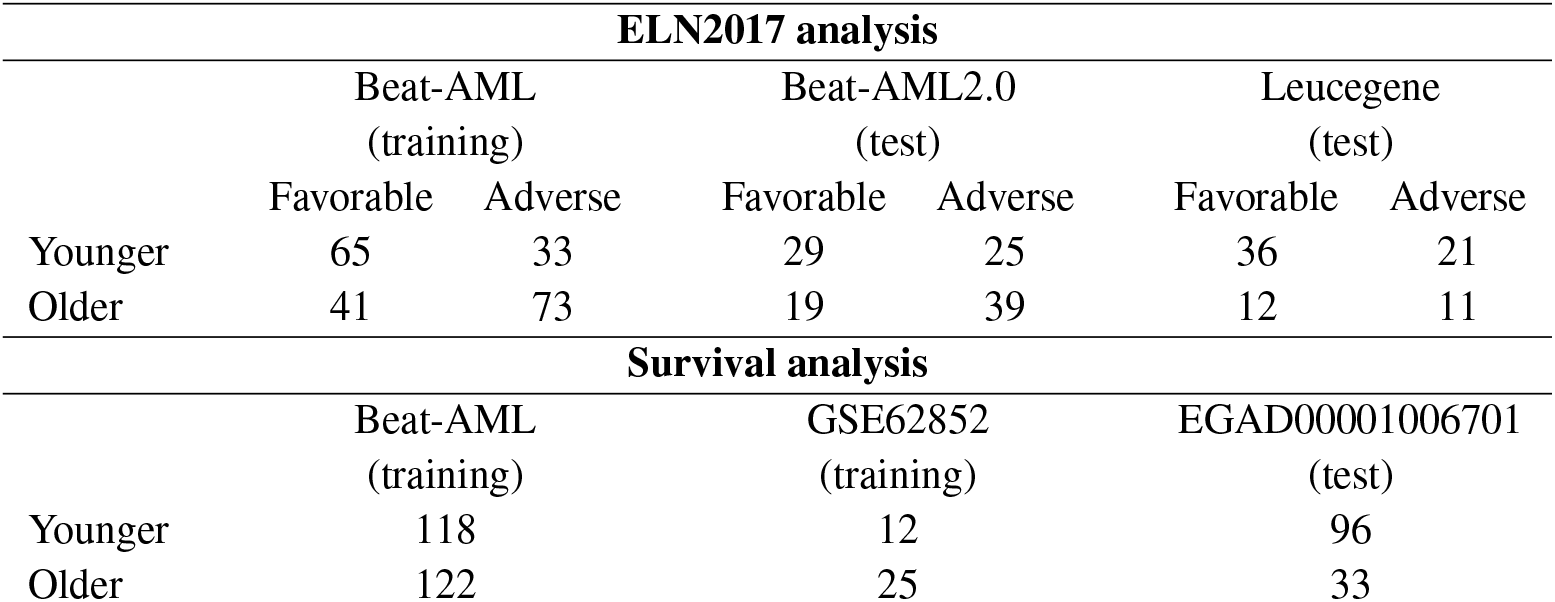
Number of transcriptome samples for ELN2017 and survival analysis. ELN2017 analysis with 404 samples and survival analysis with 406 samples.

The cohorts for the ELN2017 analysis were used in the ML process. We used Beat-AML to train the models, Beat-AML2.0 and Leucegene to test them. To avoid data leakage and to confirm the effectiveness of the evaluation, we used Beat-AML and Beat-AML2.0 as distinct groups. Thus, samples of the Beat-AML2.0 cohort that belonged to Beat-AML patients were removed from the analysis. Beat-AML and Beat-AML2.0 samples were obtained from the dbGAP database, accessions ID phs001657.v1.p1 and phs001657.v2.p1, respectively. Furthermore, we also used the Leucegene cohort (accessions ID GSE49642, GSE52656, and GSE62190) to test the models.

The cohorts were authorized for use and underwent a quality control process. We checked the quality of the raw data using fastQC version 0.11.9 ^16^ and MultiQC version 1.9 ^17^. As complementary quality control, we verified the sequencing protocol information and contamination with KmerExplor ^18^. Information about quality control, genetic variants, and clinical information for the training and test cohorts can be found in supplementary data (Additional_file1 [younger] and Additional_file2 [older]).

### Generating training k-mer count matrices

A k-mer is a substring extracted from a biological sequence (read) of fastq raw data. To extract and count k-mers from the fastq files, we used Kmtricks ^12^, a tool to count k-mers in large datasets and produce a k-mer count matrix across multiple samples. To generate the k-mer count matrices, we used only samples classified as favorable or adverse risk at the time of initial diagnosis. Samples from intermediate patients were not used due to the difficulty in defining this group in real-life ^19^: some patients fall between classes and are classified as favorable/intermediate or intermediate/adverse due to the difficulty to define risk stratification.

To avoid noisy data, we applied filters from Kmtricks and counted k-mers with a minimum abundance of 4 (i.e., a k-mer has to be found at least four times in a sample) and present in at least 5% of all samples of the analyzed cohort. We generated one matrix for younger patients and another for older patients using the Beat-AML cohort.

### Selecting k-mers

Due to the high dimensionality of the data, we applied a feature selection step to the k-mer count matrices from the training cohort. As shown in Fig. 1.A, from the k-mers count generated by Kmtricks, we applied three filters to each k-mer individually. (1) Expressed k-mer: we selected a k-mer if its count was different from zero in at least 70% of the samples in the favorable or adverse risk groups. (2) Highly expressed outliers removal: we removed k-mers with values considered outliers. A k-mer was considered an outlier if its count was higher than the third quartile across all samples. (3) Differential expression: we applied a coefficient of variation to the k-mer counts between favorable and adverse samples to assess differential expression.

**Figure 1.**
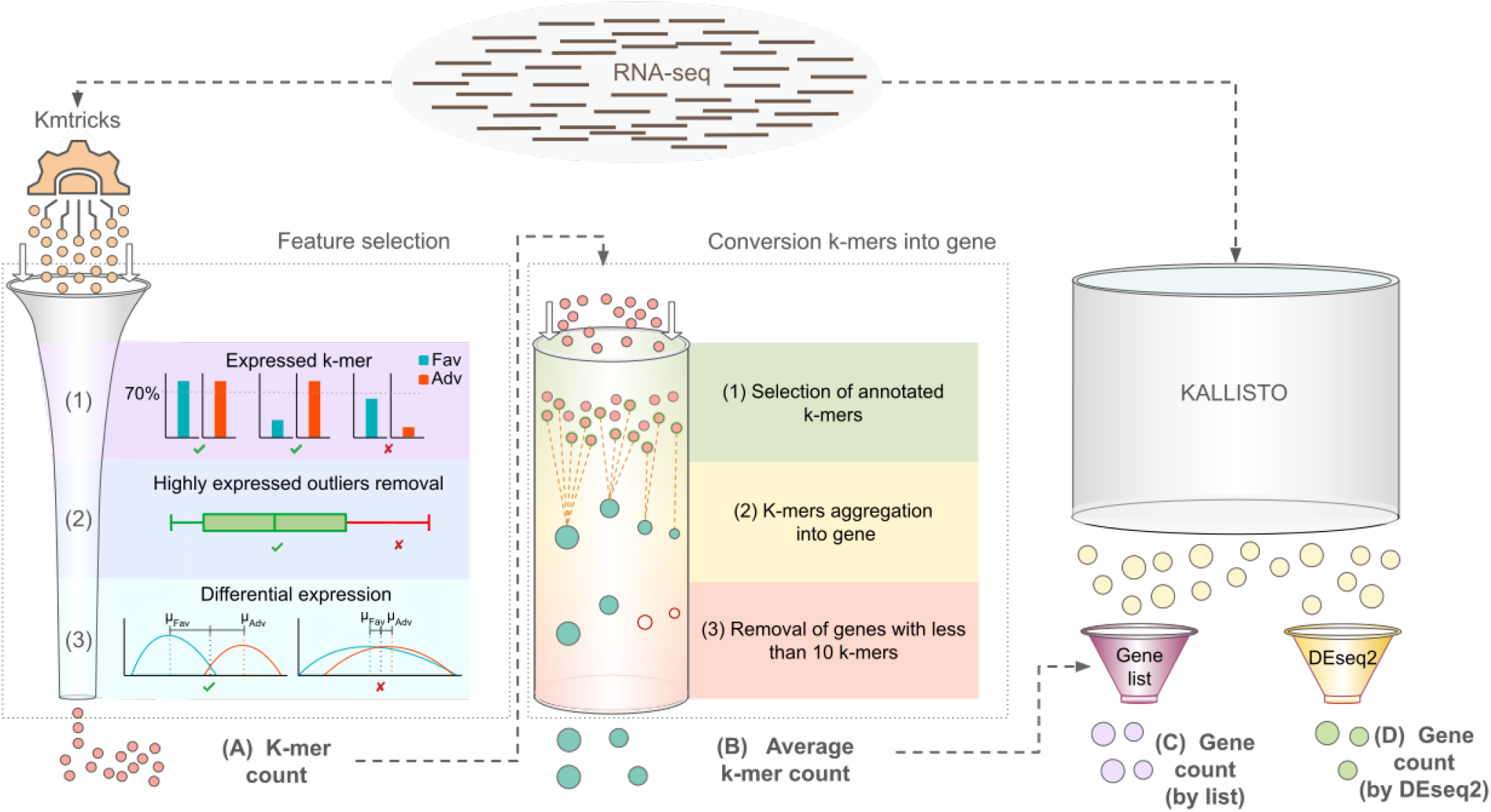
Overview to generate different counting methods. (A) K-mer count generated in the feature selection step. (B) Average k-mer count generated by the conversion from k-mers into gene. (C) Gene count generated by Kallisto and genes selected using a gene list. (D) Gene count generated by Kallisto and genes selected using DEseq2.

The coefficient of variation is defined by Equation 1.

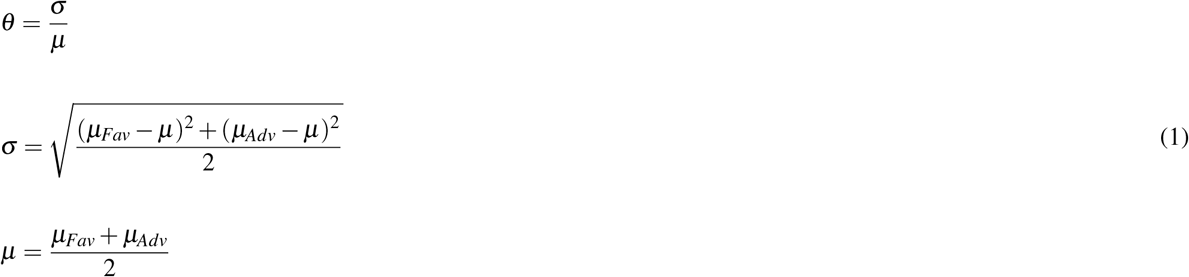

where *σ* is the standard deviation of the distances between the average k-mer count of favorable and adverse and the overall average k-mer count across all samples. *µ* is the sum of the average k-mer counts from favorable and adverse samples divided by the number of prognostic groups.

A k-mer is selected if the coefficient of variation is greater than or equal to 1. This process generated new k-mer count matrices, one for younger patients and another for older patients, which were used to train the ML models.

### Generating test k-mer count matrices

To evaluate the ML models, we needed to test k-mer count matrices using the same k-mers used in the training. To generate these k-mer count matrices, we applied “Back_to_sequences” ^20^, a tool that indexes a set of k-mers of interest and computes their occurrences in sequences (k-mer count). We provide to “Back_to_sequences” the k-mer list selected in the feature selection step and the fastq files from Beat-AML2.0 and Leucegene. “Back_to_sequences” then generated the k-mer count for each sample, allowing us to construct test k-mer count matrices for younger and older patients. Finally, k-mer counts lower than 4 in the matrices were replaced with zero to reduce noise in the data.

### Machine Learning methods

Using the k-mer count matrices with selected k-mers, we built machine learning models to predict whether a patient has favorable or adverse risk, considering both younger and older patients. We selected six Machine Learning algorithms used in other studies to predict cancer ^21,22^, including AML ^23,24^. We used three complex models: Neural Network (NN), Random Forest (RF), and eXtreme Gradient Boosting (XGB); and three less complex models: Decision Tree (DT), K-nearest neighbors (KNN), and Logistic Regression (LR).

We implemented the algorithms using the Scikit-Learn version 1.2.2 ^25^ and XGBoost version 1.7.4 ^26^ packages in Python. For each model, we applied a grid search with different parameters and a stratified cross-validation with 10-folds. The trained models were then used to predict the favorable or adverse risk in the test k-mer count matrices.

The models were evaluated by accuracy, sensitivity, specificity, and Matthew’s correlation coefficient (MCC), metrics that express the relationship between True/False Positives and True/False Negatives. True Positives (TP) are favorable samples that were correctly predicted as favorable; True Negatives (TN) are adverse samples that were predicted as adverse; False Positives (FP) are adverse samples that were wrongly predicted as favorable; and False Negatives (FN) are favorable samples that were predicted as adverse. The metrics are defined by Equations 2 to 5.

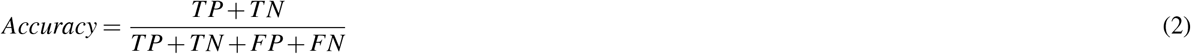

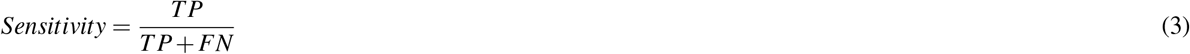

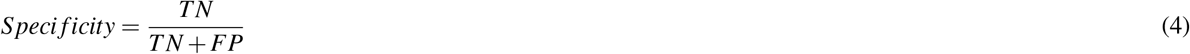

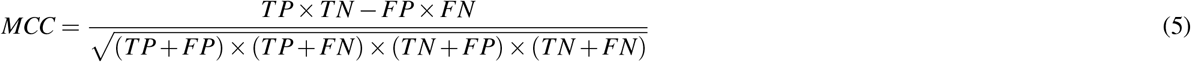

Additionally, we used the AUC (Area Under the ROC Curve), which summarizes the relationship between the True Positive Rate and the False Positive Rate into a single value.

A general overview of the steps for performing the model training and prediction is provided in Supplementary Fig. S1.

Furthermore, details of the validation step are presented in Supplementary Fig. S2.

All scripts, models, Jupyter notebooks, and data used in the machine learning analyses are available in the supplementary data.

### Mapping and annotation

We identified and associated the k-mers selected during the feature selection step with the genes they belong to. To achieve this, we applied STAR 2.7.8a ^27^ to map the k-mers to a reference human genome, the GRCh38 assembly, and we used SAMtools 1.11 ^28^ to generate flexible alignment formats (SAM and BAM files) containing mapping positions. Then, we implemented an R script using the Ensembl REST API ^29^ to request the gene annotation for each k-mer based on the SAM and BAM files. A k-mer was assigned to a gene only if it was mapped to a single position in the genome with 100% alignment.

We also had k-mers that were aligned in different positions or were not 100% aligned. In these cases, we classified them as “unannotated k-mers”. If a k-mer was not aligned, we classified it as “unmapped k-mers”.

### Expression counting methods

We used different methods to quantify the information from reads, as presented in Fig. 1: (A) k-mer count; (B) average k-mer count; (C) gene count (by list); and (D) gene count (by DEseq).

Method A uses the k-mer count directly from Kmtricks, based on the k-mers retained after the high-dimensionality reduction step (process described in “Selecting k-mers” section).

The method B is a conversion from k-mers to genes that includes three steps: (1) selection of annotated k-mers specific to each age group; (2) aggregation of k-mers into the genes that they belong to, based on average count; (3) selection of genes with more than 10 k-mers, which was indispensable considering that genes with few k-mers can be poorly representative. Additionally, only genes exclusive to each age group were retained, with genes common to multiple groups being removed to ensure group specificity.

Method C used the classic “gene count”. We used a widely used gene method, computed by Kallisto ^30^, and then, we selected the genes by a list of genes identified in the B method.

Method D also used “gene count” from Kallisto. To select the genes, we used DEseq2^31^, a known method for analyzing differential gene expression in RNA-Seq.

### Survival statistical analysis

Identification of genes impacting patients’ survival, defined as the time from diagnosis to death, was performed based on survival analysis. Two models were considered, one for younger patients (age ≤ 60 years old) and one for older patients (age > 60 years old). For each age group, we proceeded as follows: from the genes identified in the counting method B, we applied a correlation test from Caret package, where if two genes have more than 95% correlation, we removed the gene with the largest mean absolute correlation. Then, gene expression was defined as “high” if gene expression value was greater or equal to the mean, or “low” if it was lower. Age was included in the analysis as it is known to impact survival. Given the large number of genes, a two-step approach was performed. First, we performed dimension reduction by selecting genes based on Cox LASSO method^32^ using Glmnet package. The penalization parameter was determined using cross-validation and was chosen such that the deviance of the model was minimal. Then, genes with a non-zero coefficient were included in a multivariate Cox model^33^ using Survival package. Proportional hazards assumption was assessed for genes and age individually using statistical tests^34^ based on Schoenfeld residuals. The effect size for each gene and age was estimated using the hazard ratio together with its 95% confidence interval.

In order to evaluate the predictive performances of selected genes and age, we compared our survival models with survival models trained using ELN risk classification as prognosis factor for both age groups. We used an independent cohort (EGAD00001006701) to obtain survival prediction and we computed for each model Harrell’s concordance-index (c-index)^35^, which is a statistic used in survival analysis to evaluate the ability to discriminate risk within a population. A c-index ranges from 0 to 1, with a value of 0.5 corresponding to a model performing as good as a random classifier and a value of 1 corresponding to a perfect discrimination.

The analysis was performed in R and can be found in supplementary data.

### Biological context

We investigated the biological context of the samples by analyzing the percentage of blast cells, mutation profile, fusion gene presence, and ratio of immune and stromal cells. We analyzed bone marrow (BM) and peripheral blood (PB) samples from the Beat-AML cohort. The blast percentage was provided by the Vizome website. Mutation and fusion gene information was obtained from metadata on cbioportal. For the mutation profile, we considered mutations in the DNMT3, TET2, IDH1, IDH2, and ASLX1 genes, which are frequently associated with clonal hematopoiesis, as described by^7^. The fusion genes considered included CBFB-MYH11, DEK-NUP214, GATA2-MECOM, MLLT3-KMT2A, PML-RARA, and RUNX1-RUNX1T1.

In order to count the immune and stromal cells, we used a previously reported method based also on k-mer counting. We designed specific k-mers from the gene list of MCP-counter ^36^ using Kmerator ^18^. Kmerator is a tool capable of generating k-mers that are unique to the requested genes (specific k-mers). We grouped the genes (averaging the k-mers) by cell type, including B lineage, CD8 T cells, T cells, cytotoxic lymphocytes, natural killer (NK) cells, dendritic, monocytic, neutrophils, fibroblasts, and endothelial cells.

We used g:Profiler ^37^ to perform Gene Ontology (GO) analysis on our list of identified genes. g:Profile search for statistically significant Gene Ontology (GO) terms, pathways, and other gene function-related terms.

Additionally, we sought to find information about the selected k-mers that did not have a corresponding gene (unannotated and unmapped k-mers). To achieve this, we searched these k-mers from younger and older patients in RJunBase^38^, a database which references transcripts splice information and associated cancer metadata at the genome scale. For the identified k-mers, we applied a univariate Cox model to estimate their impact on survival, selecting only those with a p-value *<* 0.05. Finally, for the significant k-mers, we also used RJunBase to verify whether they were associated with tumors.

## Results

### Predicting favorable and adverse outcome patients

Using Kmtricks, we generated k-mer matrices from 98 younger and 114 older patients, containing 364,846,009 and 399,034,012 k-mers, respectively. Feature selection reduced these to 35,098 k-mers (younger) and 63,929 k-mers (older). These reduced matrices were used with six machine learning (ML) models to classify patients into favorable or adverse ELN risk groups using count method A (k-mer counts): Decision Tree (DT), K-nearest Neighbors (KNN), Logistic Regression (LR), Neural Network (NN), Random Forest (RF), and eXtreme Gradient Boosting (XGB).

Among younger patients, LR yielded the highest accuracy (92%), while XGB performed best in older patients (91%) (Fig. 2.A1). UMAP projections (Fig. 2.A2) demonstrated clear separation between favorable and adverse groups, supporting the biological relevance of selected k-mers.

**Figure 2.**
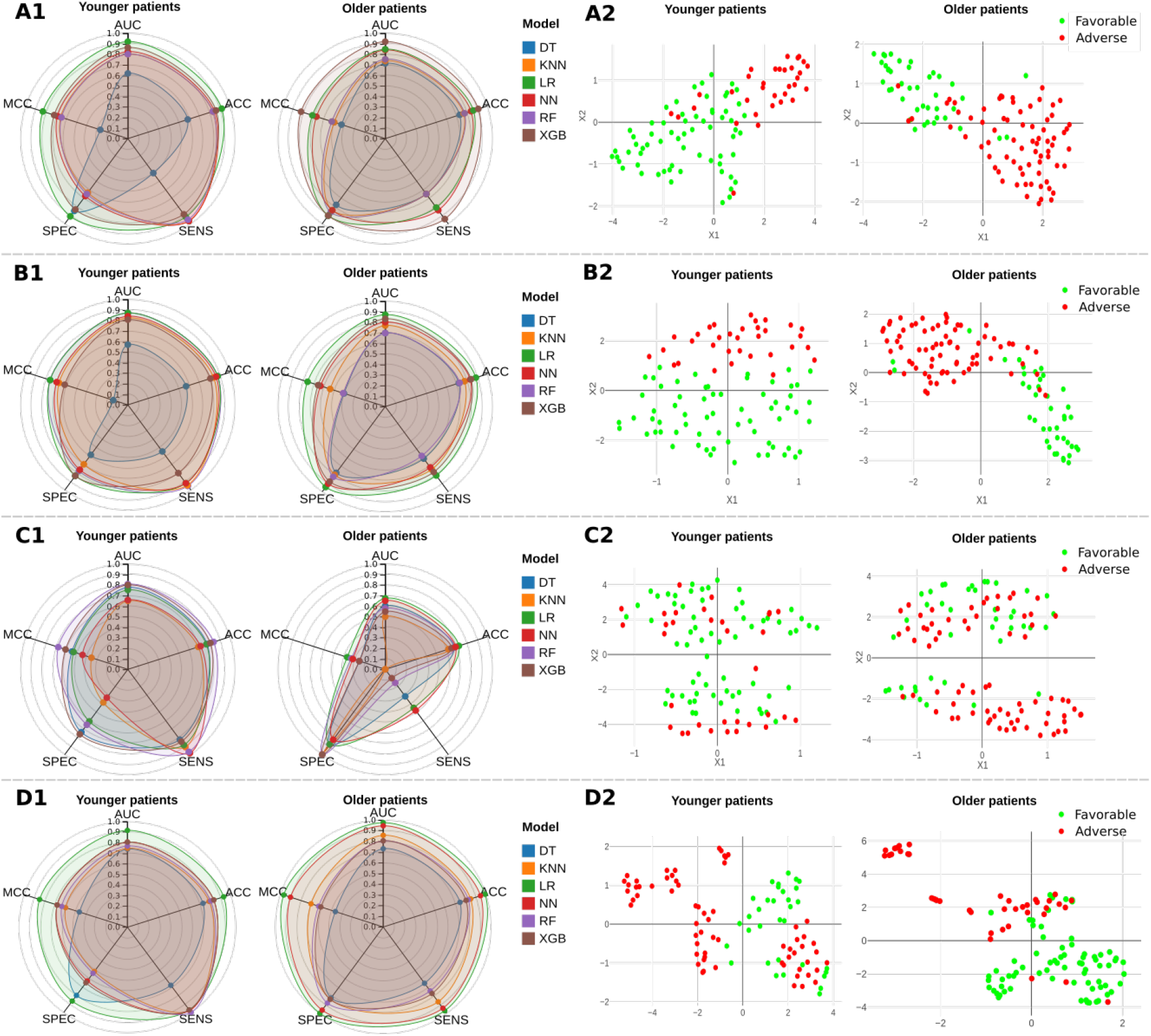
Predicting favorable and adverse risk with k-mers (A1), average k-mer (B1), and genes selected by list (C1), genes selected by DEseq2 (D) counts using Decision Tree (DT), K-nearest neighbors (KNN), and Logistic Regression (LR), Neural Network (NN), Random Forest (RF), and eXtreme Gradient Boosting (XGB) models. Metrics Area under the curve (AUC), accuracy (ACC), sensitivity (SENS), specificity (SPEC), and Matthew’s correlation coefficient (MCC) for evaluating models in younger and older patients. UMAP projection with k-mer (A2), average k-mer (B2), gene selected by list (C2), and genes selected by DEseq2 (D) counts.

Using count method B (average k-mer counts converted to gene-level features), we identified 99 genes for younger and 250 genes for older patients (supplementary Tables S1 and S2). Since the predictions with the six models using the count method A were well-performed, we trained then six models with method B. LR and RF achieved the highest accuracy in younger patients (88%), while LR remained optimal for older patients (89%) (Fig. 2.B1). UMAP projections continued to show strong separation (Fig. 2.B2).

Using count method C, we quantified the same genes with Kallisto. Although RF performed best in younger patients, its accuracy dropped by 6% relative to method B (Fig. 2.C1). In older patients, LR achieved only 67% accuracy. UMAP projections (Fig. 2.C2) reflected this decline, with less clear group separation.

With count method D, we applied DESeq2 to Kallisto-quantified data from 30,352 genes. Differential expression analysis (adjusted *p* ≤ 0.05) yielded 2,736 genes for younger and 5,538 for older patients. LR achieved the highest performance in both younger (93%) and older (99%) groups (Fig. 2.D1). However, UMAP projections (Fig. 2.D2) showed poor separation in younger patients and only modest improvement in older patients. Overall, k-mer-based methods yielded comparable or superior predictions to traditional gene quantification, particularly in the younger cohort.

Complete performance metrics, including validation on Beat-AML2.0 and Leucegene cohorts, are provided in Supplementary Tables S3–S6.

### Survival in younger and older patients

Identification of genes impacting patients’ survival was performed for 99 genes for younger and 250 genes for older groups from Beat-AML and the GSE62852 cohorts.

We first analyzed the correlation between genes, where we removed 11 genes highly correlated in the younger group, and 31 genes in the older group. After we applied a dimension reduction step, 10 variables were retained by the Cox LASSO for the younger group, including age. For the older group, age was the only variable selected confirming the difference observed in the transcriptome in this group compared to younger patients. The proportional hazards assumption was verified for the final Cox models including the selected variables (see Supplementary Tables S7-S10). The results of the models are shown in Figure 3.A. For younger patients, SLC29A2 and GLCCI1 expressions have p-values lower than 0.05 (p = 0.0001 and 0.0263, respectively), making them significant risk factors (Fig. A). Additionally, their hazard ratios (HR) are greater than 1 (HR = 4.38 [2.07-9.27] and 2.48 [1.11-5.53], respectively), indicating that when these k-mers are expressed, the instantaneous risk of death is higher compared to when they are not. Furthermore, LINGO3 expression shows borderline significance (p = 0.0718), with an HR greater than 1 (HR = 1.94 [0.94-3.98]). For both young and old patients, age has a significant impact on survival (p = 0.0163 and p < 0.0001, respectively). This means that each additional year of age is associated with an increased instantaneous risk of death (HR = 1.03 [1.01-1.06] and 1.06 [1.04-1.09], respectively).

**Figure 3.**
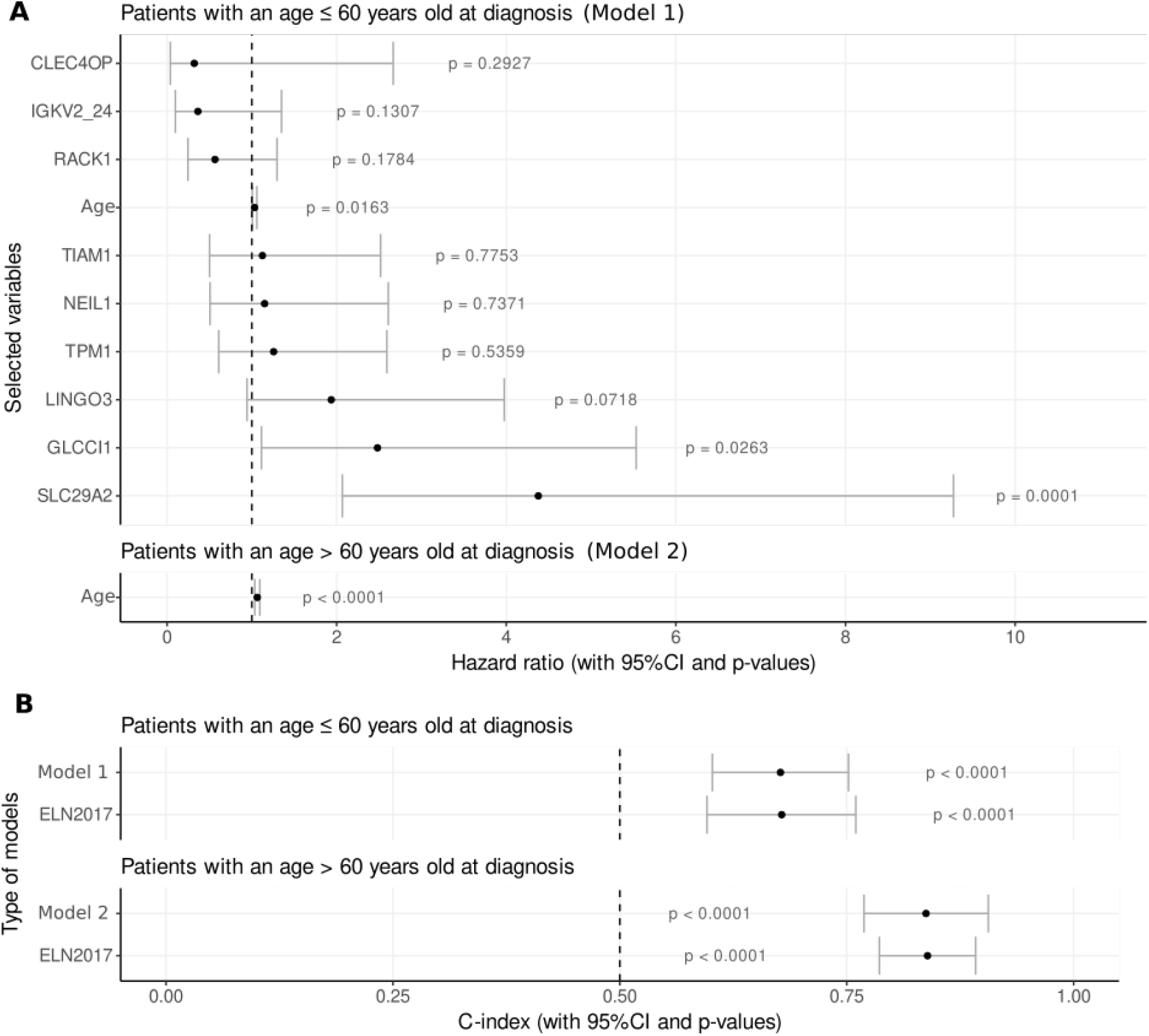
Death prediction for young (below 60 years old) and old patients (above 60 years old) using survival analysis (Cox proportional hazards (PH) models). (A) Hazard ratio (HR) of features impacting patient survival, previously selected from k-mer expressions and age using a Cox-Lasso approach, for patients below 60 years old (model 1) and above 60 years old (model 2). (B) Comparative performance of death predictions using the C-index of Cox PH models, comparing the ELN2017 score with a panel of selected k-mer expressions and age (for young patients), and age only (for old patients) as prognostic factors. p-values for C-index from z-test against 0.5.

Using the c-index, we compared our models with survival models using ELN2017 status as a prognostic factor. The results were similar for both younger and older patients (c-index = 0.677 [0.602-0.752] and 0.678 [0.596-0.760], respectively, for younger patients; 0.837 [0.769-0.906] and 0.839 [0.786-0.892], respectively, for older patients) (Fig. 3.B).

### The complexity of transcriptomic profiles in older patients

To further explore transcriptomic differences between age groups, we conducted a cross-prediction analysis using the average k-mer count data (counting method B) (Fig. 4.A). In this test, we reversed the application of trained models: the six machine learning models trained on younger patients were used to predict risk in older patients, and vice versa.

**Fig. 4.**
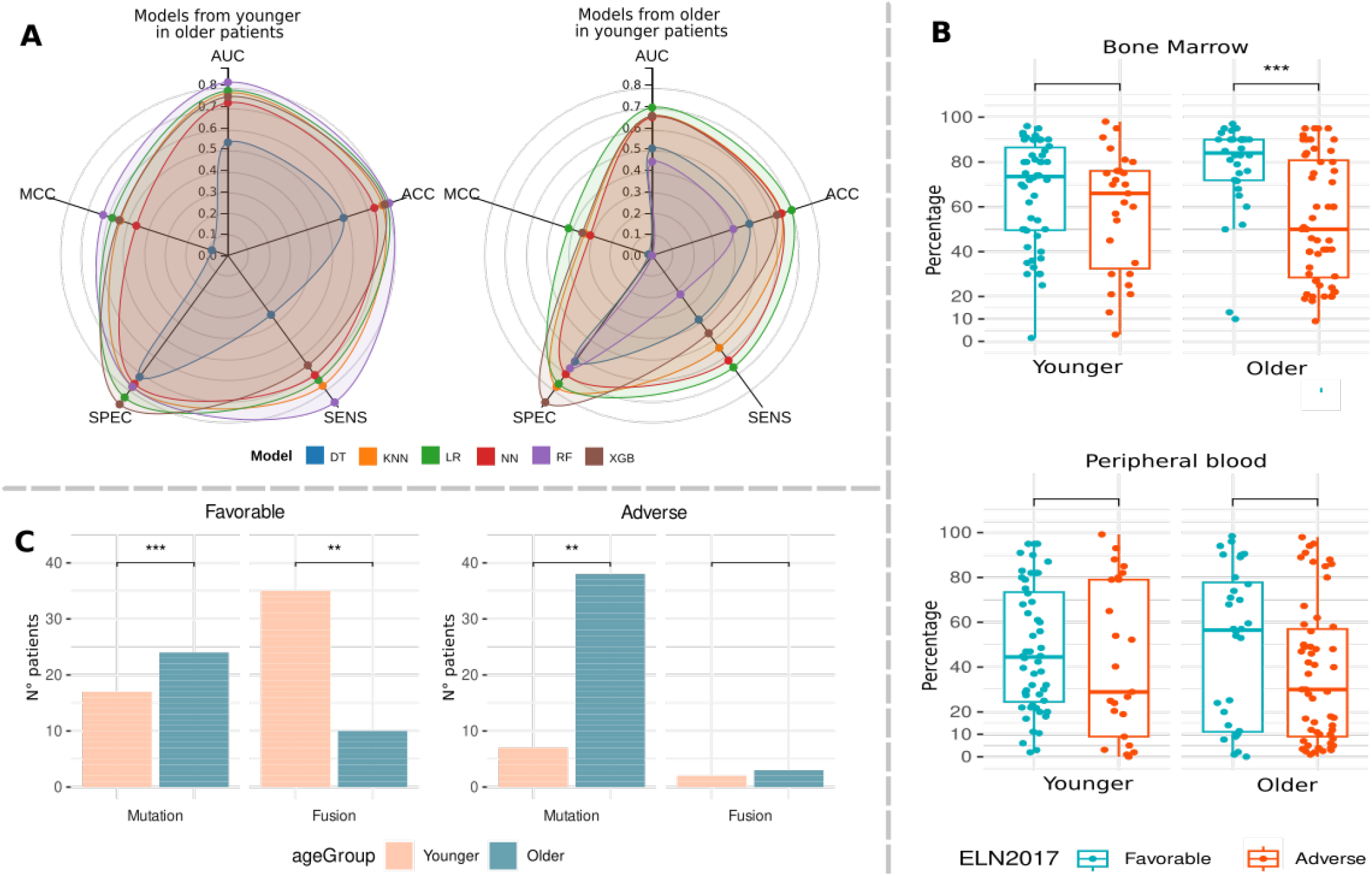
(A) Performance for models from younger patients to predict in older patients and models from older patients to predict in younger patients. (B) Percent of blast cells in bone marrow and peripheral blood samples. (C) Number of younger and older patients with mutations and fusion genes in favorable and adverse risk. p-values in (B) and (C) from Wilcoxon test: ‘****’: *p* ≤ 0.0001; ‘***’: *p* ≤ 0.001; ‘**’: *p* ≤ 0.01; ‘*’: *p* ≤ 0.05; ‘‘: *p* > 0.05.

The models trained on younger patients performed reasonably well when applied to older patients, with the random forest (RF) model achieving an accuracy of 81%. This represented only an 8% drop compared to the performance of the same model trained and tested on older patients (89% accuracy).

In contrast, models trained on older patients struggled to predict risk in younger patients. The best-performing model in this setting was logistic regression (LR), which reached only 70% accuracy. This result reflected a more substantial performance decline of 18% compared to the model trained and applied within the younger cohort (89% accuracy).

These findings suggest that transcriptomic patterns in older patients differ substantially from those in younger individuals. The decreased cross-predictive performance may indicate that aging has a significant impact on k-mer/gene expression, leading to greater transcriptomic complexity and variability.

To investigate possible biological explanations for these differences, we proposed some hypotheses regarding the biological and cellular content that might underlie this discrepancy. These differences could arise from intrinsic tumor cell profiles, external factors such as the types of cells present in the tumor microenvironment, or physiological parameters associated with aging. To evaluate these possibilities, we first examined the percentage of blast cells in both bone marrow (BM) and peripheral blood (PB) samples (Fig. 4.B). In older patients, we observed a significant difference in BM blast percentage between favorable and adverse risk groups. This difference was not significant in the younger group and, as expected, was not observed in PB samples.

We then assessed molecular profiles of tumor cells by comparing the presence of mutations in genes frequently associated with clonal hematopoiesis in older patients, as well as the presence of fusion genes, which are typically more frequent in younger patients. As expected, significant differences were observed between the older and younger groups (Fig. 4.C). The highest mutation rates were seen in older patients with adverse risk, whereas fusion genes were most prevalent in younger patients with favorable risk. Together, these findings reinforce the notion of distinct tumor behavior patterns and genomic alterations associated with aging.

Next, we evaluated the composition of the bone marrow microenvironment by profiling immune and stromal cell markers. In younger patients, only dendritic cells showed significant differences between favorable and adverse risk groups. In contrast, older patients with adverse risk exhibited significantly higher levels of B and T lymphocytes, as well as endothelial and fibroblast cells (Fig. 5). These observations point to a broader reshaping of the microenvironment in older patients, potentially contributing to disease progression and therapy resistance. When analyzing peripheral blood, this difference persisted only for endothelial cells in older patients (see Supplementary Fig. S3). Moreover, the alterations observed in the bone marrow microenvironment were further supported by the analysis of gene counts across the same groups (see Supplementary Fig. S4).

**Figure 5.**
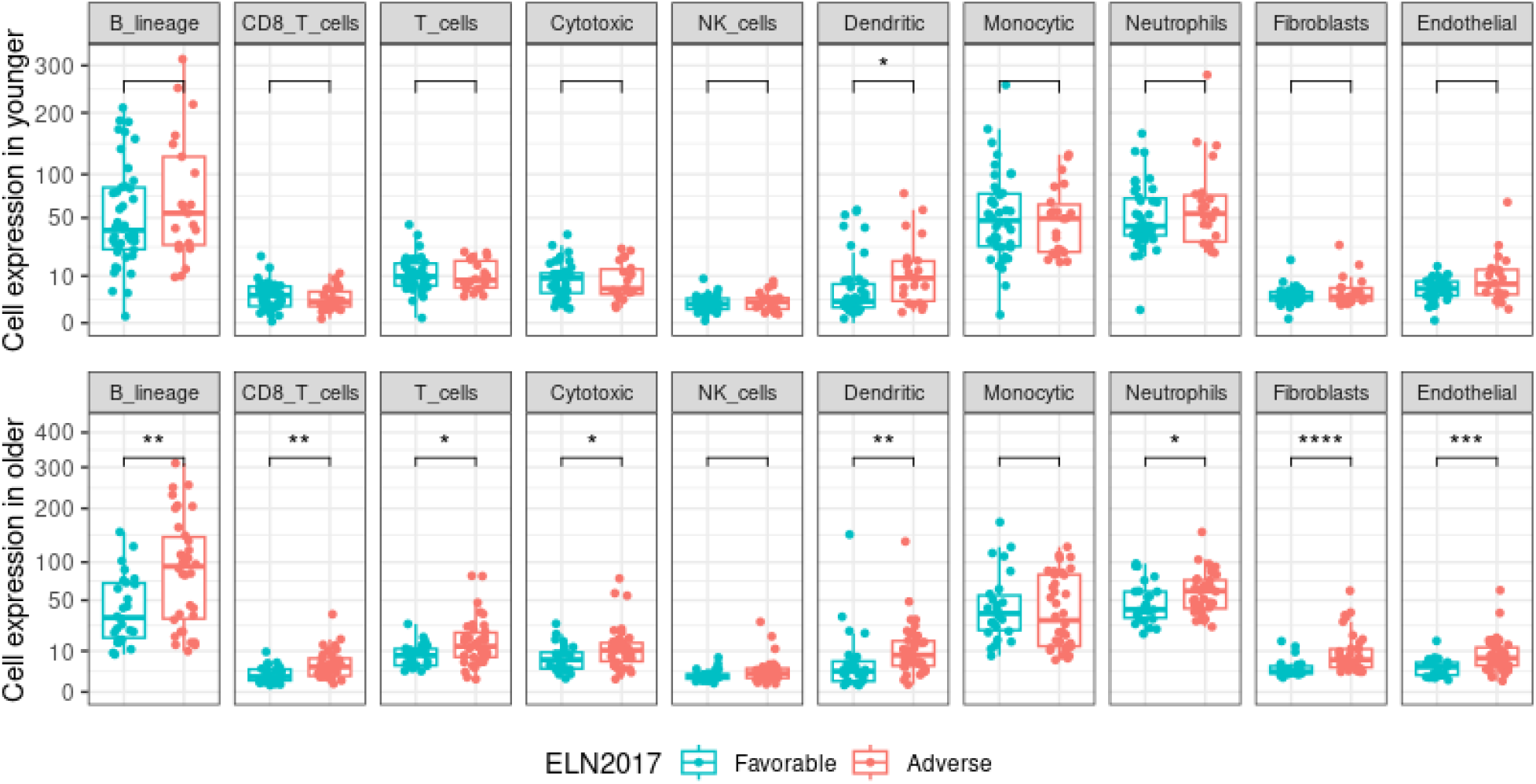
Expression of immune and stromal cells in bone marrow (BM) samples and comparison of favorable and adverse risk (Wilcoxon test). ‘****’: *p* ≤ 0.0001; ‘***’: *p* ≤ 0.001; ‘**’: *p* ≤ 0.01; ‘*’: *p* ≤ 0.05; ‘‘: *p >* 0.05.

We also performed a Gene Ontology (GO) enrichment analysis on the list of younger and older identified genes, which revealed functional categories uniquely enriched in the transcriptome profiles of older patients. These included immune-related processes (such as peptide antigen binding, antigen processing, and components of the MHC protein complex) as well as stromal cell-associated functions (including cell adhesion and cardiac fibroblast cell development). These functional enrichments are consistent with the immune and stromal cell populations identified in the bone marrow microenvironment (Supplementary Fig. S5).

### Analysis of unannotated and unmapped k-mers

To explore transcriptomic regions not captured by gene annotations, we analyzed the set of unannotated and unmapped k-mers. This analysis revealed 8,951 such k-mers in younger patients and 8,478 in older patients, as shown in the Table 2. To investigate whether these sequences represented splicing events, we queried them against the RJunBase database. This approach identified 219 candidate splice junctions in the younger cohort and 572 in the older cohort.

**Table 2.** Identification of spliced junctions in younger and older patients using RJunBase.

|  | Unannotated and unmapped | RJun splicing | Impact on survival | Tumor associated |
| --- | --- | --- | --- | --- |
| Younger | 8,951 | 219 | 148 | 0 |
| Older | 8,478 | 572 | 303 | 12 |

We next assessed the association between these candidate junctions and patient survival. Based on statistical significance (p < 0.05), 148 k-mers were selected in younger patients and 303 in older patients. Interestingly, despite the number of splice junction candidates initially detected, none in the younger group remained associated with known annotated splice junctions after filtering. In contrast, 12 splice junctions were retained in the older group following survival analysis, indicating a more prominent contribution of splicing alterations to transcriptomic variability and prognosis in older AML patients.

The 12 splicing junctions identified in older patients belong to the genes E2F2 (1 splicing), TAL1 (1 splicing), PAWR (5 splicing), SETBP1 (1 splicing), NEDD4L (1 splicing), FHL2 (1 splicing), and TIMP3 (2 splicing). Among them, TAL1 is an oncogene of the bHLH transcription factor aberrantly expressed in 60% of cases of T-cell acute lymphoblastic leukemia^39^, while mutations in SETBP1 have been identified in various hematologic malignancies, including acute myeloid leukemia (AML)^40^. The differential expression of TAL1 and SETBP1 splicing junctions are presented in the Supplementary material (Fig. S6).

These results suggest that splicing events captured by unannotated and unmapped k-mers, particularly in older patients, may contribute to disease biology and could serve as novel prognostic markers or therapeutic targets.

## Discussion

In this study, we applied machine learning models to transcriptome data using a k-mer based approach to investigate the differences in the risk stratification of younger and older AML patients. Using RNA-seq data, we analyzed transcriptome expression levels through different counting methods to detect qualitative and quantitative changes in specific conditions. We observed that k-mer proved to be a valuable approach to investigate RNA-seq data due to two main reasons. First, it allowed us to analyze the data on a large scale without pre-mapping or assembly. Second, it captures the biological information more precisely, including single mutations^11^ and splicing junctions.

Our study results showed that genes identified using counting method B (average k-mer count) were capable of distinguishing favorable from adverse risk. We found 99 genes for younger patients and 250 genes for older patients. These genes were identified based on k-mers selected using ELN risk information, without any other previous knowledge. The good performance of the ML prediction confirmed the efficiency of both the feature selection step (counting method A) and the gene to k-mer conversion step (counting method B) in predicting favorable and adverse risks. When comparing our ELN risk prediction based on k-mers with the prediction performed using gene quantification that considered whole-gene reads (counting method C), we obtained the best results with our method, showing that k-mers contain sufficient information and that full-length transcript information is not required.

When performing survival analysis to assess time to death in the young group, four variables had a significant impact: SLC29A2, GLCCI1, LINGO3, and age. Interestingly, SLC29A2, GLCCI1, and LINGO3 are three genes associated with tumor resistance or progression. SLC29A2 is a nucleoside transporter involved in treatment resistance in cancers and AML^41,42^. The GLCCI1 gene, which encodes a protein of unknown function, has recently been described as a binding partner of the DYRK1A kinase, and functional evidence supports DYRK1A as a potential tumor suppressor involved in chemoresistance in AML^43,44^. LINGO3 has been described as a metastatic biomarker in cancer^45^. Additionally, LINGO3 has been implicated in CRISPR screens using the MOLM13 AML cell line in response to venetoclax, contributing to increased drug resistance^46^.

In contrast, age emerged as the sole significant variable influencing survival in older patients. This finding aligns with previous reports, including the study by Straube et al.^47^, which emphasized the dominant role of age in AML prognosis and its influence on the ELN classification. The limited influence of transcriptomic features on survival in older patients may reflect a broader heterogeneity in disease biology and patient physiology in this population.

To explore the underlying biological differences between age groups, we assessed the bone marrow (BM) microenvironment. In older patients with adverse risk, we observed a reduced blast cell fraction, suggesting infiltration or expansion of non-leukemic cell types. This prompted a deeper investigation into the immune and stromal cell landscape of BM samples. Stromal components, particularly endothelial and fibroblast cells, were found in higher abundance in older adverse-risk patients. These cell types are known to contribute to leukemia pathogenesis by modulating the tumor microenvironment. For example, endothelial cells can support leukemic stem cell maintenance and mediate interactions with immune cells^48^, while fibroblasts promote leukemic cell survival via extracellular matrix remodeling and cytokine secretion^49^. Our findings are consistent with reports suggesting that the aging bone marrow niche becomes increasingly permissive to leukemic progression^50^.

Additionally, our results showed that immune cells are also highly expressed in adverse older patients. The impact on these patients can be explained by the fact that the immune system undergoes profound changes with aging and immune cells are shown to support leukemogenesis and resistance to therapy^51^. A high proportion of regulatory T cells was already reported as of poor prognosis by interfering with immunologic synapse formation^52^. Also, regulatory B cells were reported with high expression in patients with poor prognosis^53^.

In summary, our results point in the same direction as the recent literature showing that leukemic cells influence the BM microenvironment to support their survival^54,55^, and may transcriptionally resemble normal immune cells^56^. Moreover, our results highlight the influence of AML cells in the BM microenvironment mainly in older patients when the risk increases, which can implicate leukemia cell survival, and also resistance to therapy in this age group.

## Data Availability

The data generated during the study are available from the corresponding author upon reasonable request.

https://osf.io/kthvb/.

## Data availability

The data analyzed in this study are publicly available in the dbGAP database (https://www.ncbi.nlm.nih.gov/gap/) with the accessions ID phs001657.v1.p1 and phs001657.v2.p1, and the GEO database (https://www.ncbi.nlm.nih.gov/geo/) accessions ID GSE49642, GSE52656, GSE62190, and GSE62852. We also used EGAD00001006701 available in European Genome-phenome Archive (EGA) (https://ega-archive.org/). The data generated during the study are available from the corresponding author upon reasonable request.

## Acknowledgements

This work has been supported by La Ligue Contre le Cancer and the Agence Nationale de la Recherche (TranSipedia and FullRNA projects).

## Author contributions statement

RS wrote the manuscript, analyzed the data, and developed the methodology. CR interpreted the data and prepared the Figure 1 and 3. MA performed the survival analysis. JR revised the manuscript and analyzed the spliced junctions. CS are generated the data for immune and stromal cells analysis. CB performed classical differential gene expression analysis. BG and AB worked to manage the AML cohorts. FR and NG interpreted the annotation. TC designed the study.

## Additional information

Supplementary data can be found at https://osf.io/kthvb/.

