## Supplementary material for "Acute myeloid leukemia risk stratification in younger and older patients through transcriptomic machine learning models"

#### Supplementary Figure S1

The pipeline for generating ML models and predicting prognosis from RNA-seq data is shown in Figure S1. From an RNA-seq cohort, we generate a training k-mer count table using Kmtricks. We then apply a feature selection step to identify k-mers relevant for distinguishing prognosis, and generate a new training k-mer count table containing only the selected k-mers. This table is used to train the ML models. For a new patient, we extract the counts of the selected k-mers from the RNA-seq data using BackToSequences and generate a test k-mer count table. This table is then used by the trained model to predict prognosis.

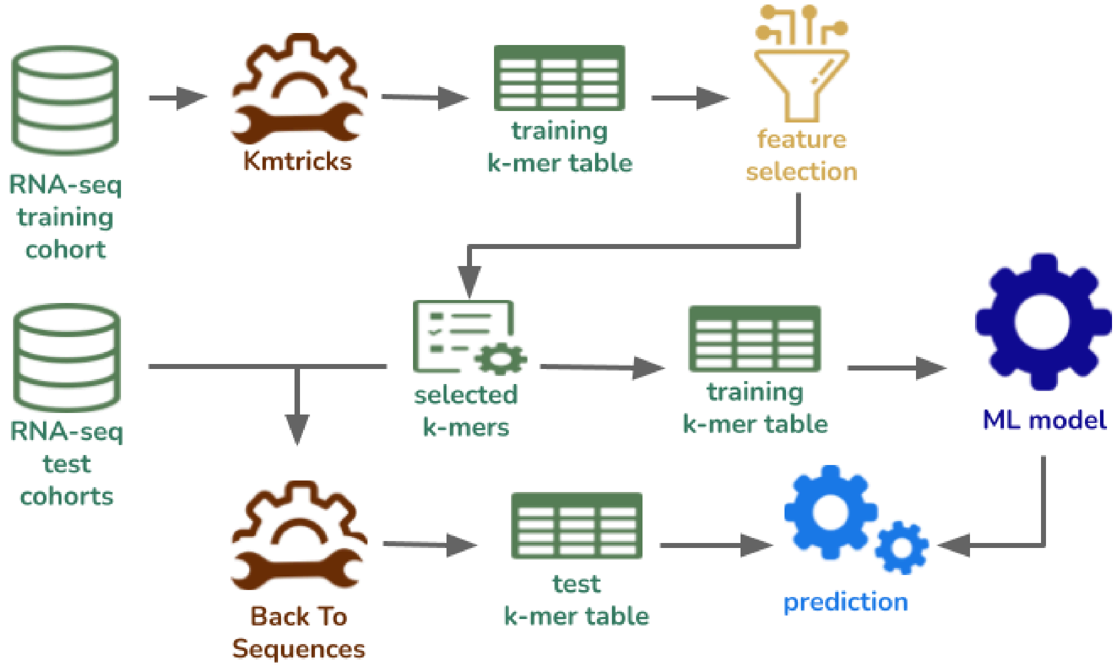


Figure S1. Pipeline to training and prediction for RNA-seq data.

**Supplementary Figure S2**

As an initial test (validation step), we analyzed the prediction if the patient has a favorable or adverse risk in younger and older patients using a k-mer approach with ML, we performed initial tests with 70% of the Beat-AML cohort to train and 30% to validate. We trained using Decision Tree (DT), K-nearest neighbors (KNN), and Logistic Regression (LR), Neural Network (NN), Random Forest (RF), and eXtreme Gradient Boosting (XGB) models. The metrics to evaluate the initial models can be seen below.

| **Younger** | | | | | |
|---|---|---|---|---|---|
| **Model** | **Accuracy** | **AUC** | **Sensitivity** | **Specificity** | **MCC** |
| DT | 0.63 | 0.53 | 0.81 | 0.25 | 0.07 |
| KNN | 0.84 | 0.75 | 1 | 0.5 | 0.64 |
| LR | 0.87 | 0.84 | 0.92 | 0.75 | 0.69 |
| NN | 0.89 | 0.86 | 0.96 | 0.75 | 0.75 |
| RF | 0.84 | 0.75 | 1 | 0.5 | 0.64 |
| XGB | 0.79 | 0.78 | 0.81 | 0.75 | 0.54 |
| **Older** | | | | | |
| **Model** | **Accuracy** | **AUC** | **Sensitivity** | **Specificity** | **MCC** |
| DT | 0.79 | 0.77 | 0.69 | 0.86 | 0.56 |
| KNN | 0.82 | 0.83 | 0.85 | 0.81 | 0.64 |
| LR | 0.88 | 0.89 | 0.92 | 0.86 | 0.76 |
| NN | 0.85 | 0.88 | 1 | 0.76 | 0.74 |
| RF | 0.85 | 0.87 | 0.92 | 0.81 | 0.71 |
| XGB | 0.94 | 0.95 | 1 | 0.9 | 0.89 |

Figure S2. Predicting favorable and adverse risk with initial models: Decision Tree (DT), K-nearest neighbors (KNN), and Logistic Regression (LR), Neural Network (NN), Random Forest (RF), and eXtreme Gradient Boosting (XGB) models. Performance of models in younger and older patients.

The best performance in younger patients was achieved by the Neural Network (NN) model achieving an accuracy of 89%. In older patients, eXtreme Gradient Boosting (XGB) achieved the best performance in all the metrics, with an accuracy of 94%.

### Supplementary Figure S3

Similarly as presented in the paper, we designed specific k-mers from the gene list of MCP-counter using Kmerator. Then, we regrouped the genes (average of k-mers) by cell type to see the expression of each cell in peripheral blood samples for younger and older patients. Natural killer (NK) cells, Fibroblasts, and Endothelial cells showed a difference between favorable and adverse in younger patients. In older, only Endothelial cells showed to be different between the risks.

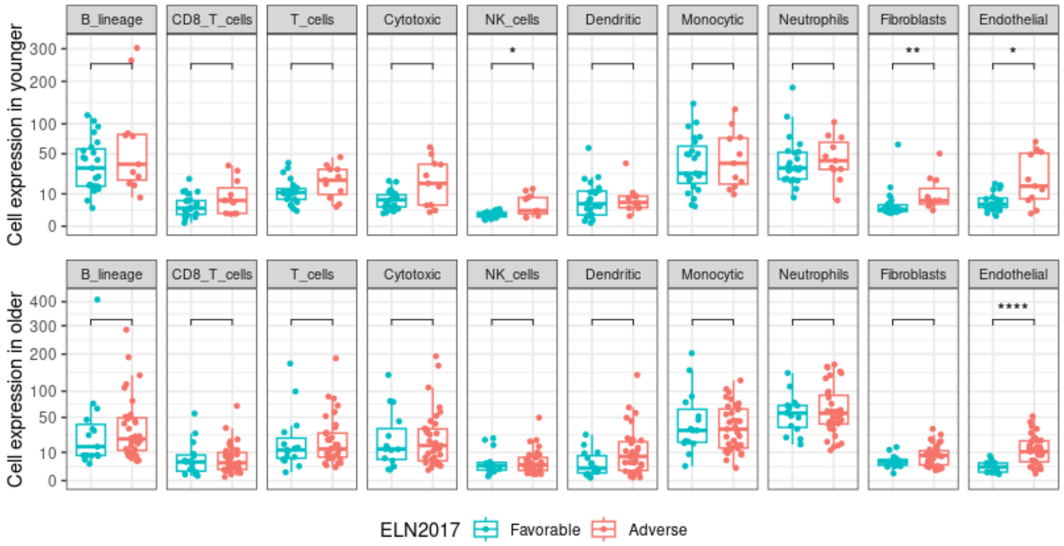


Figure S3. Expression of immune and stromal cells in peripheral blood (PB) samples and comparison of favorable and adverse risk (Wilcoxon test). '****': p <= 0.0001; '***': p <= 0.001 ; '**': p <= 0.01; '*': p <= 0.05; ' ': p > 0.05.

**Supplementary Figure S4**

To validate the immune and stromal cell abundance results previously obtained through k-mer counting (using the MCP-counter gene list with Kmerator), we performed an additional analysis based on gene counts generated by Kallisto (method D, as described in the manuscript). We applied xCell to generate cell abundance profiles from these gene counts. xCell is a machine learning-based method that estimates cell infiltration in tissues from transcriptomic data, providing relative enrichment scores that reflect the proportion of each cell type within the total cellular composition of the sample. The boxplots (Fig. S4 A, B, C, D) represent the distribution of xCell enrichment scores for the indicated cell types within the corresponding subgroups. These estimated relative abundances allow us to confirm differences in the proportions of cell types between younger and older patients.

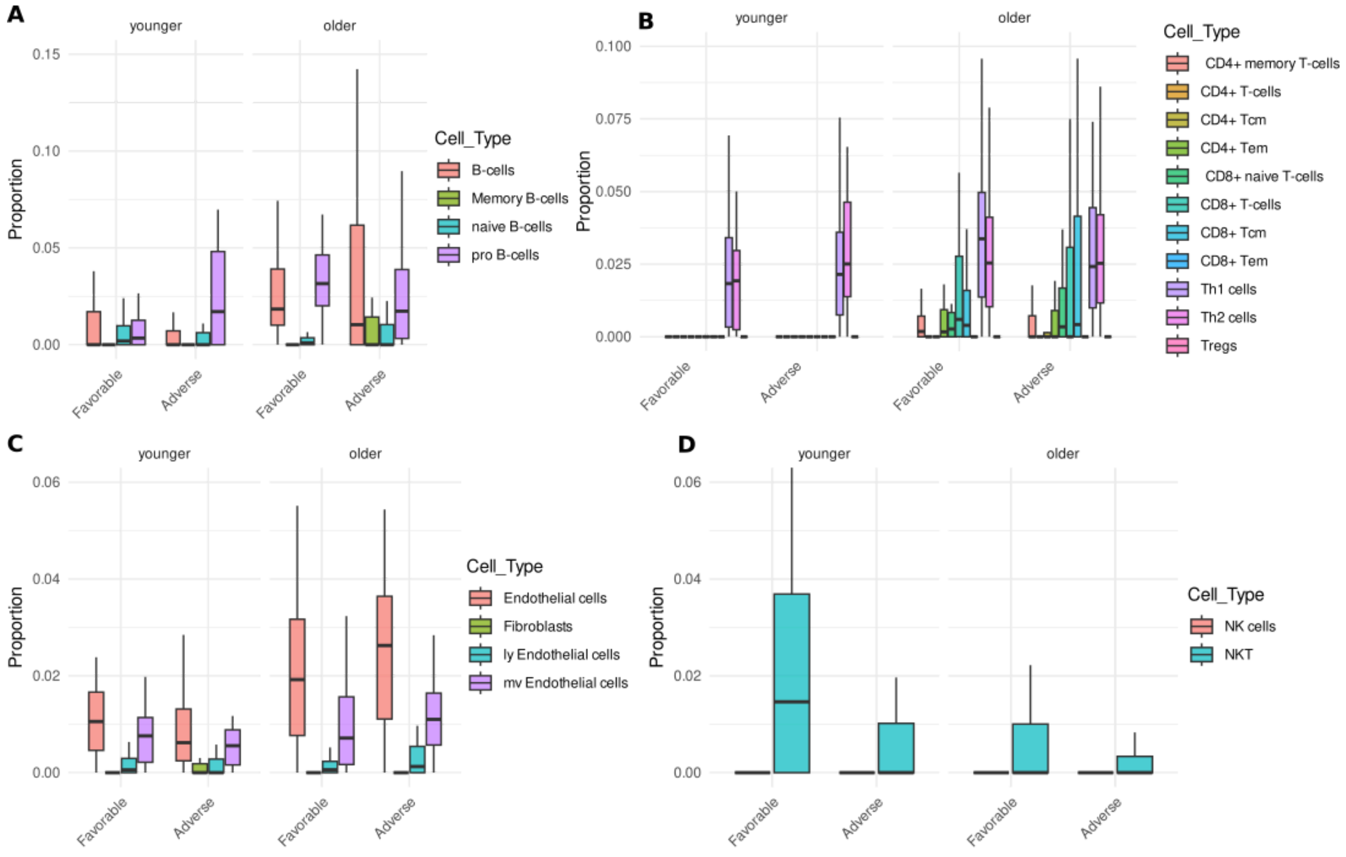


Figure S4. Proportion of B-cells (A), proportion of T-cells (B), proportion of stromal cells (C), and proportion of natural killer (NK) cells (D) in younger and older by risk.

**Supplementary Figure S5**

We used g:Profiler to explore the biological interactions of the genes identified by count method B. g:Profile finds statistically significant Gene Ontology (GO) terms, pathways, and other gene function-related terms.

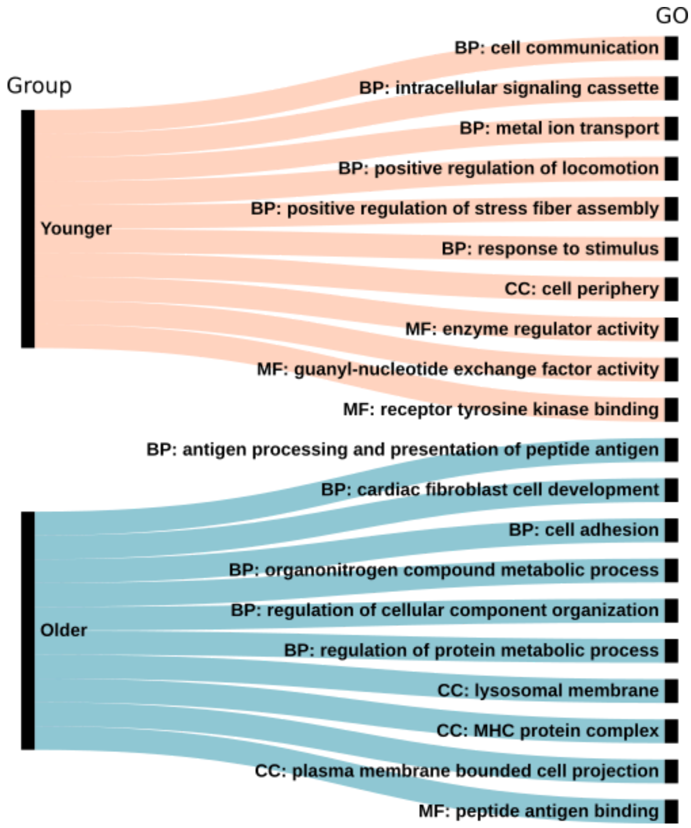


Figure S5. Gene Ontology (GO) for younger and older groups. Molecular function (MF), biological process (BP), or cellular component (CC).

Figure S5 shows the analysis of the enrichment in gene ontology (GO) in the set of genes from younger and older patients. Interestingly, specific functions were only observed in the transcriptome data of older patients in link with the immune response (peptide antigen binding, antigen processing, and MHC protein complex) as well as stromal cell features (cell adhesion, cardiac fibroblast cell development) and correlated with observed cell populations.

**Supplementary Figure S6**

Expression of TAL1_LS004 and SETBP1_LS007 splicing in younger and older patients (Fig. S6.A,B) and expression of samples in [RJunBase](RJunBase) (Fig.S6.C,D).

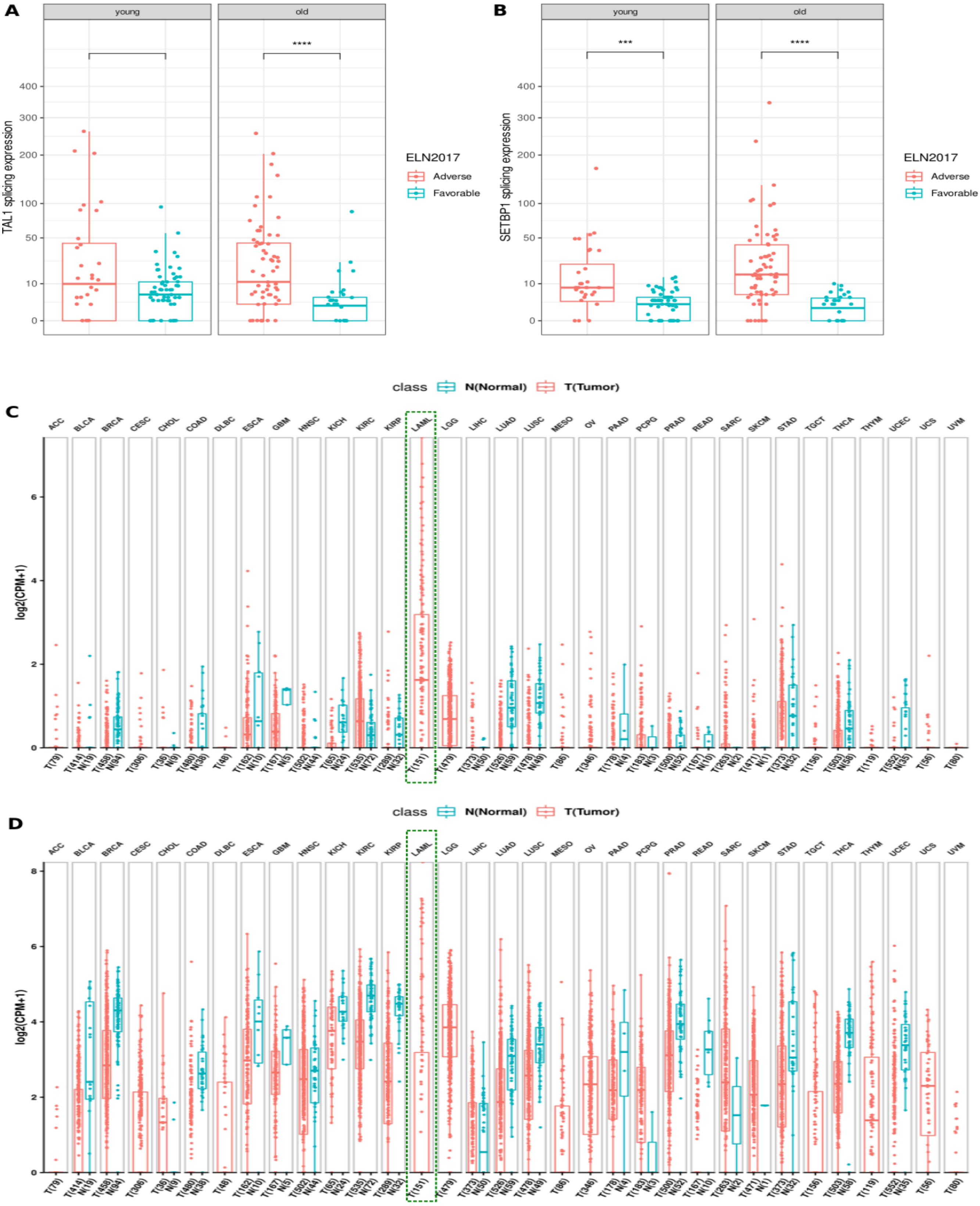


Figure S6. K-mer expression of TAL1_LS004 splicing (A), SETBP1_LS007 splicing (B) Expression of TAL1_LS004 (C) and SETBP1_LS007 (D) in the Cancer Genome Atlas (TCGA) samples in the “Summary” tab on the" Linear Splice Junction Detail“ page of TAL1_LS004 and SETBP1_LS007.

### Supplementary Table S1. Genes identified for younger patients

| Gene | Selected k-mers |
|---|---|
| ABR | 15 |
| ACVR2B | 12 |
| AMOT | 13 |
| ANKS6 | 434 |
| ATP1A3 | 14 |
| B4GAT1-DT | 25 |
| BAIAP2L2 | 14 |
| BCAS4 | 28 |
| BMI1 | 21 |
| BMP8B | 58 |
| CD44 | 30 |
| CD96 | 47 |
| CEP192 | 18 |
| CEP72 | 13 |
| CLCN5 | 85 |
| CLDND2 | 12 |
| CLEC11A | 61 |
| CLEC4OP | 24 |
| CLEC9A | 18 |
| COL8A2 | 20 |
| COMMD3-BMI1 | 23 |
| CPNE8 | 12 |
| CRACR2A | 11 |
| CROCC2 | 22 |
| CRTC3 | 20 |
| CYP2U1 | 12 |
| CYP2U1-AS1 | 12 |
| DGKH | 38 |
| DNMT3B | 12 |
| EHMT1 | 13 |
| EPN1 | 17 |
| F2R | 12 |
| FAM86B3P | 28 |
| FARP2 | 31 |
| FBN1 | 20 |
| GAPDH | 11 |
| GLCCI1 | 27 |
| GPR84-AS1 | 50 |
| HIVEP3 | 12 |
| IGKV2-23 | 11 |
| IGKV2-24 | 12 |
| INPP5B | 21 |
| KHDC1 | 13 |
| KIAA1549 | 37 |
| KLRG1 | 12 |
| LAMA5 | 275 |
| LINC02811 | 12 |
| LINC02891 | 32 |
| LINGO3 | 14 |
| LPO | 261 |

| | |
|---|---|
| MALAT1 | 12 |
| MAMDC2 | 14 |
| MAMDC2-AS1 | 12 |
| MGST1 | 31 |
| MPO | 5502 |
| MRPL42 | 17 |
| NEIL1 | 39 |
| PAQR6 | 13 |
| PFKM | 18 |
| PI4KA | 12 |
| PIF1 | 12 |
| PLXNA1 | 54 |
| PPIE | 58 |
| PPP1R9B | 11 |
| QRFP | 35 |
| RACK1 | 13 |
| RBFOX2 | 91 |
| RPS6KA1 | 12 |
| SAMD11 | 39 |
| SCAMP5 | 14 |
| SH3PXD2B | 42 |
| SLC16A1-AS1 | 52 |
| SLC25A23 | 52 |
| SLC29A2 | 25 |
| SNORD116-4 | 15 |
| SPATS2L | 11 |
| STK25 | 21 |
| STXBP5 | 29 |
| TALAM1 | 12 |
| TBC1D29P | 44 |
| TBCD | 13 |
| TIAM1 | 48 |
| TMEM213 | 22 |
| TMEM273 | 37 |
| TPM1 | 35 |
| TPPP | 13 |
| TRIP10 | 19 |
| TRO | 16 |
| TSPAN14-AS1 | 35 |
| TUBG2 | 16 |
| TXNIP | 85 |
| UICLM | 22 |
| VSTM1 | 17 |
| XKR6 | 14 |
| XYLB | 14 |
| ZG16B | 71 |
| ZHX3 | 30 |
| ZNF263 | 12 |
| ZNF532 | 91 |

### Supplementary Table S2. Genes identified for older patients

| Gene | Selected k-mers |
|---|---|
| AAK1 | 13 |
| ABCB1 | 55 |
| ABCG1 | 71 |
| ABLIM1 | 20 |
| ADCY3 | 28 |
| AGBL2 | 17 |
| AMPD3 | 22 |
| ANPEP | 12 |
| APOL3 | 53 |
| ARAP2 | 28 |
| ARHGEF10 | 11 |
| ARHGEF17 | 12 |
| ARMCX5-GPRASP2 | 14 |
| ATP10A | 218 |
| ATP8B3 | 35 |
| ATP8B4 | 23 |
| BAALC | 190 |
| BAALC-AS1 | 198 |
| BAALC-AS2 | 78 |
| BACH1-IT1 | 12 |
| BCL2L14 | 23 |
| BIN1 | 36 |
| BMAL1 | 16 |
| BRF1 | 23 |
| C16orf46 | 35 |
| C16orf74 | 17 |
| C2orf88 | 67 |
| C8orf88 | 22 |
| CA5B | 56 |
| CAB39L | 18 |
| CALN1 | 20 |
| CASK | 11 |
| CAT | 148 |
| CATSPER2 | 16 |
| CCDC136 | 15 |
| CCDC24 | 32 |
| CD34 | 115 |
| CD74 | 11 |
| CDC42BPA | 62 |
| CELSR1 | 36 |
| CIITA | 40 |
| CNGB3 | 33 |
| COBLL1 | 48 |
| COL24A1 | 53 |

| | |
|---|---|
| COL6A2 | 61 |
| CRELD2 | 18 |
| CRHBP | 40 |
| CRNDE | 140 |
| CRYBG3 | 11 |
| CTTN | 20 |
| CUL4A | 12 |
| CYFIP2 | 17 |
| DAZAP2 | 14 |
| DGAT1 | 11 |
| DGKE | 11 |
| DIPK1B | 19 |
| DLGAP3 | 29 |
| DMXL2 | 206 |
| DTWD2 | 31 |
| DUSP1 | 17 |
| EFNB1 | 11 |
| EGFL7 | 44 |
| EIF1B-AS1 | 14 |
| ELOVL7 | 33 |
| EPHA1-AS1 | 20 |
| ERG | 13 |
| ESAM | 24 |
| F2RL1 | 186 |
| FAM124B | 24 |
| FAM30A | 14 |
| FANK1 | 12 |
| FBLN5 | 13 |
| FBXO22 | 16 |
| FGFBP2 | 22 |
| FILNC1 | 11 |
| FIS1 | 12 |
| FLJ12825 | 13 |
| FOXO1 | 17 |
| GATA3 | 35 |
| GBP4 | 18 |
| GBP5 | 15 |
| GGTA1 | 12 |
| GNAI1 | 29 |
| GNG11 | 15 |
| GNL3 | 11 |
| GPR34 | 11 |
| GPRASP1 | 14 |
| GPRC5B | 34 |
| GRAP2 | 13 |

| | |
|---|---|
| GYPC | 302 |
| H1-0 | 14 |
| H2BC4 | 11 |
| HAL | 19 |
| HIBCH | 67 |
| HILPDA-AS1 | 36 |
| HLA-B | 20 |
| HLA-DPA1 | 28 |
| HLA-DPB1 | 33 |
| HLA-DRA | 27 |
| HOMER3 | 14 |
| HPGD | 14 |
| HSD17B4 | 23 |
| IFI16 | 37 |
| IFI44 | 77 |
| IFT140 | 11 |
| IGF2BP3 | 15 |
| IL12RB2 | 52 |
| IL6ST | 19 |
| INHBA | 17 |
| IPO9-AS1 | 162 |
| IQCG | 54 |
| IQCK | 64 |
| IQSEC2 | 11 |
| ISG20 | 49 |
| JUN | 20 |
| KANK1 | 17 |
| KBTBD11-OT1 | 11 |
| KCNA3 | 20 |
| KIAA2026 | 13 |
| LAPTM5 | 43 |
| LDOC1 | 76 |
| LGMN | 62 |
| LINC00174 | 14 |
| LINC00467 | 11 |
| LINC01237 | 12 |
| LINC02147 | 26 |
| LINC02573 | 15 |
| LINC02767 | 35 |
| LMLN | 72 |
| LRP6 | 30 |
| LTB | 14 |
| MAN1A1 | 50 |
| MAP6D1 | 16 |
| MCOLN2 | 14 |

| | |
|---|---|
| MEF2C-AS2 | 11 |
| MIAT | 71 |
| MIR130AHG | 20 |
| MITF | 20 |
| MLANA | 13 |
| MLYCD | 12 |
| MMP19 | 144 |
| MMP28 | 28 |
| MN1 | 510 |
| MON1A | 14 |
| MREG | 63 |
| MRPL33 | 19 |
| MTAP | 64 |
| MX1 | 80 |
| MYRIP | 14 |
| NAV1 | 163 |
| NECAB1 | 17 |
| NEDD4L | 78 |
| NEK11 | 12 |
| NHSL1 | 39 |
| NOXRED1 | 16 |
| NPR3 | 88 |
| NREP | 21 |
| NRG4 | 24 |
| OCEL1 | 11 |
| OIP5 | 25 |
| OIP5-AS1 | 26 |
| OTULINL | 23 |
| PAK1 | 38 |
| PAWR | 74 |
| PBX3 | 53 |
| PEAR1 | 30 |
| PHKA2-AS1 | 11 |
| PIWIL4 | 95 |
| PIWIL4-AS1 | 95 |
| PPHLN1 | 22 |
| PPP1R26 | 11 |
| PREP | 20 |
| PREPL | 17 |
| PRICKLE1 | 49 |
| PRKCH | 132 |
| PROM1 | 100 |
| PRR5 | 24 |
| PRR5L | 55 |
| PSMB8 | 15 |

| | |
|---|---|
| PSMB9 | 37 |
| PTGFRN | 50 |
| PXDC1 | 16 |
| QPRT | 13 |
| RAB8A | 34 |
| RABGAP1L | 17 |
| RAMP1 | 18 |
| RBKS | 18 |
| RBPMS-AS1 | 13 |
| RDX | 197 |
| RFX8 | 20 |
| RIPOR3 | 187 |
| RNF130 | 21 |
| RNF217 | 111 |
| ROBO4 | 117 |
| RORA | 20 |
| RORA-AS1 | 20 |
| RTN1 | 12 |
| SAMD15 | 11 |
| SCARF1 | 70 |
| SCD | 27 |
| SCN1B | 14 |
| SCRN1 | 93 |
| SERPINB6 | 125 |
| SHANK3 | 510 |
| SIAE | 17 |
| SLC2A9 | 29 |
| SLC37A3 | 86 |
| SLC39A11 | 13 |
| SLC39A8 | 14 |
| SLC41A3 | 31 |
| SLC6A9 | 32 |
| SMAGP | 14 |
| SNX33 | 11 |
| SPAG16 | 65 |
| SPRY1 | 12 |
| ST3GAL3 | 54 |
| STARD4-AS1 | 17 |
| STOM | 71 |
| SUSD3 | 30 |
| SV2A | 32 |
| SYN3 | 73 |
| SYNE2 | 15 |
| TAL1 | 38 |
| TAP1 | 37 |

| | |
|---|---|
| TBCK | 15 |
| TBXAS1 | 17 |
| TC2N | 24 |
| TGFBR3 | 20 |
| TICAM2-AS1 | 30 |
| TIMP3 | 73 |
| TMC4 | 23 |
| TMC6 | 16 |
| TMC8 | 12 |
| TMED7-TICAM2 | 30 |
| TMEM204 | 11 |
| TP53I11 | 37 |
| TRH | 41 |
| TRPS1 | 264 |
| TSPAN18 | 37 |
| TSPAN7 | 30 |
| USP54 | 15 |
| VSIG2 | 12 |
| YEATS2 | 23 |
| YEATS2-AS1 | 23 |
| YPEL4 | 20 |
| ZBP1 | 79 |
| ZC3H12C | 195 |
| ZNF277-AS1 | 14 |
| ZNF626 | 13 |
| ZNF662 | 185 |

### Supplementary Table S3. Metrics Method A

| Younger - Beat-AML2.0 and Leucegene | | | | | |
|---|---|---|---|---|---|
| **Model** | **Accuracy** | **AUC** | **Sensitivity** | **Specificity** | **MCC** |
| DT | 0.59 | 0.62 | 0.4 | 0.85 | 0.27 |
| KNN | 0.83 | 0.8 | 0.97 | 0.63 | 0.66 |
| LR | 0.92 | 0.92 | 0.92 | 0.91 | 0.83 |
| NN | 0.85 | 0.82 | 0.97 | 0.67 | 0.69 |
| RF | 0.83 | 0.8 | 0.95 | 0.65 | 0.65 |
| XGB | 0.86 | 0.86 | 0.89 | 0.83 | 0.72 |
| **Younger - Beat-AML2.0** | | | | | |
| **Model** | **Accuracy** | **AUC** | **Sensitivity** | **Specificity** | **MCC** |
| DT | 0.72 | 0.73 | 0.66 | 0.8 | 0.46 |
| KNN | 0.83 | 0.82 | 1 | 0.64 | 0.7 |
| LR | 0.94 | 0.94 | 1 | 0.88 | 0.89 |
| NN | 0.85 | 0.84 | 1 | 0.68 | 0.73 |
| RF | 0.83 | 0.82 | 0.97 | 0.68 | 0.68 |
| XGB | 0.85 | 0.85 | 0.86 | 0.84 | 0.7 |
| **Younger - Leucegene** | | | | | |
| **Model** | **Accuracy** | **AUC** | **Sensitivity** | **Specificity** | **MCC** |
| DT | 0.46 | 0.55 | 0.19 | 0.9 | 0.13 |
| KNN | 0.82 | 0.78 | 0.94 | 0.62 | 0.62 |
| LR | 0.89 | 0.91 | 0.86 | 0.95 | 0.79 |
| NN | 0.84 | 0.81 | 0.94 | 0.67 | 0.66 |
| RF | 0.82 | 0.78 | 0.94 | 0.62 | 0.62 |
| XGB | 0.88 | 0.86 | 0.92 | 0.81 | 0.73 |
| **Older - Beat-AML2.0 and Leucegene** | | | | | |
| **Model** | **Accuracy** | **AUC** | **Sensitivity** | **Specificity** | **MCC** |
| DT | 0.73 | 0.71 | 0.65 | 0.78 | 0.43 |
| KNN | 0.77 | 0.74 | 0.65 | 0.84 | 0.5 |
| LR | 0.86 | 0.85 | 0.81 | 0.9 | 0.71 |
| NN | 0.84 | 0.84 | 0.84 | 0.84 | 0.67 |
| RF | 0.78 | 0.75 | 0.65 | 0.86 | 0.52 |
| XGB | 0.91 | 0.92 | 0.94 | 0.9 | 0.82 |
| **Older - Beat-AML2.0** | | | | | |
| **Model** | **Accuracy** | **AUC** | **Sensitivity** | **Specificity** | **MCC** |
| DT | 0.72 | 0.73 | 0.66 | 0.8 | 0.46 |
| KNN | 0.83 | 0.82 | 1 | 0.64 | 0.7 |
| LR | 0.94 | 0.94 | 1 | 0.88 | 0.89 |
| NN | 0.85 | 0.84 | 1 | 0.68 | 0.73 |
| RF | 0.83 | 0.82 | 0.97 | 0.68 | 0.68 |
| XGB | 0.85 | 0.85 | 0.86 | 0.84 | 0.7 |
| **Older - Leucegene** | | | | | |
| **Model** | **Accuracy** | **AUC** | **Sensitivity** | **Specificity** | **MCC** |
| DT | 0.61 | 0.62 | 0.42 | 0.82 | 0.25 |
| KNN | 0.74 | 0.75 | 0.5 | 1 | 0.57 |
| LR | 0.87 | 0.88 | 0.75 | 1 | 0.77 |
| NN | 0.78 | 0.78 | 0.83 | 0.73 | 0.56 |
| RF | 0.52 | 0.53 | 0.33 | 0.73 | 0.07 |
| XGB | 0.87 | 0.86 | 1 | 0.73 | 0.76 |

### Supplementary Table S4. Metrics Method B

| Younger - Beat-AML2.0 and Leucegene | | | | | |
|---|---|---|---|---|---|
| **Model** | **Accuracy** | **AUC** | **Sensitivity** | **Specificity** | **MCC** |
| DT | 0.57 | 0.57 | 0.55 | 0.59 | 0.14 |
| KNN | 0.84 | 0.82 | 0.94 | 0.7 | 0.67 |
| LR | 0.88 | 0.87 | 0.92 | 0.83 | 0.76 |
| NN | 0.86 | 0.84 | 0.92 | 0.76 | 0.7 |
| RF | 0.88 | 0.87 | 0.95 | 0.78 | 0.76 |
| XGB | 0.81 | 0.81 | 0.8 | 0.83 | 0.62 |
| **Younger - Beat-AML2.0** | | | | | |
| **Model** | **Accuracy** | **AUC** | **Sensitivity** | **Specificity** | **MCC** |
| DT | 0.52 | 0.51 | 0.62 | 0.4 | 0.02 |
| KNN | 0.85 | 0.84 | 0.97 | 0.72 | 0.72 |
| LR | 0.87 | 0.87 | 0.93 | 0.8 | 0.74 |
| NN | 0.81 | 0.8 | 0.97 | 0.64 | 0.65 |
| RF | 0.87 | 0.86 | 1 | 0.72 | 0.76 |
| XGB | 0.81 | 0.82 | 0.79 | 0.84 | 0.63 |
| **Younger - Leucegene** | | | | | |
| **Model** | **Accuracy** | **AUC** | **Sensitivity** | **Specificity** | **MCC** |
| DT | 0.61 | 0.65 | 0.5 | 0.81 | 0.31 |
| KNN | 0.82 | 0.79 | 0.92 | 0.67 | 0.62 |
| LR | 0.89 | 0.89 | 0.92 | 0.86 | 0.77 |
| NN | 0.89 | 0.9 | 0.89 | 0.9 | 0.78 |
| RF | 0.89 | 0.89 | 0.92 | 0.86 | 0.77 |
| XGB | 0.81 | 0.81 | 0.81 | 0.81 | 0.6 |
| **Older - Beat-AML2.0 and Leucegene** | | | | | |
| **Model** | **Accuracy** | **AUC** | **Sensitivity** | **Specificity** | **MCC** |
| DT | 0.72 | 0.7 | 0.61 | 0.78 | 0.4 |
| KNN | 0.78 | 0.77 | 0.74 | 0.8 | 0.54 |
| LR | 0.89 | 0.87 | 0.81 | 0.94 | 0.76 |
| NN | 0.83 | 0.8 | 0.71 | 0.9 | 0.63 |
| RF | 0.73 | 0.7 | 0.58 | 0.82 | 0.41 |
| XGB | 0.84 | 0.83 | 0.77 | 0.88 | 0.66 |
| **Older - Beat-AML2.0** | | | | | |
| **Model** | **Accuracy** | **AUC** | **Sensitivity** | **Specificity** | **MCC** |
| DT | 0.83 | 0.83 | 0.84 | 0.82 | 0.64 |
| KNN | 0.79 | 0.82 | 0.89 | 0.74 | 0.6 |
| LR | 0.88 | 0.86 | 0.79 | 0.92 | 0.72 |
| NN | 0.9 | 0.88 | 0.84 | 0.92 | 0.77 |
| RF | 0.83 | 0.8 | 0.74 | 0.87 | 0.61 |
| XGB | 0.91 | 0.9 | 0.84 | 0.95 | 0.8 |
| **Older - Leucegene** | | | | | |
| **Model** | **Accuracy** | **AUC** | **Sensitivity** | **Specificity** | **MCC** |
| DT | 0.43 | 0.44 | 0.25 | 0.64 | -0.12 |
| KNN | 0.74 | 0.75 | 0.5 | 1 | 0.57 |
| LR | 0.91 | 0.92 | 0.83 | 1 | 0.84 |
| NN | 0.65 | 0.66 | 0.5 | 0.82 | 0.33 |
| RF | 0.48 | 0.48 | 0.33 | 0.64 | -0.03 |
| XGB | 0.65 | 0.65 | 0.67 | 0.64 | 0.3 |

### Supplementary Table S5. Metrics Method C

| | | | | | |
|---|---|---|---|---|---|
| **Younger - Beat-AML2.0 and Leucegene** | | | | | |
| **Model** | **Accuracy** | **AUC** | **Sensitivity** | **Specificity** | **MCC** |
| DT | 0.78 | 0.78 | 0.82 | 0.74 | 0.55 |
| KNN | 0.69 | 0.65 | 0.91 | 0.39 | 0.36 |
| LR | 0.77 | 0.75 | 0.89 | 0.61 | 0.53 |
| NN | 0.71 | 0.66 | 0.98 | 0.33 | 0.44 |
| RF | 0.84 | 0.81 | 0.97 | 0.65 | 0.68 |
| XGB | 0.81 | 0.8 | 0.85 | 0.76 | 0.61 |
| **Younger - Beat-AML2.0** | | | | | |
| **Model** | **Accuracy** | **AUC** | **Sensitivity** | **Specificity** | **MCC** |
| DT | 0.78 | 0.78 | 0.79 | 0.76 | 0.55 |
| KNN | 0.69 | 0.68 | 0.79 | 0.56 | 0.36 |
| LR | 0.78 | 0.77 | 0.86 | 0.68 | 0.55 |
| NN | 0.7 | 0.68 | 0.97 | 0.4 | 0.45 |
| RF | 0.87 | 0.86 | 1 | 0.72 | 0.76 |
| XGB | 0.8 | 0.8 | 0.76 | 0.84 | 0.6 |
| **Younger - Leucegene** | | | | | |
| **Model** | **Accuracy** | **AUC** | **Sensitivity** | **Specificity** | **MCC** |
| DT | 0.79 | 0.77 | 0.83 | 0.71 | 0.55 |
| KNN | 0.7 | 0.6 | 1 | 0.19 | 0.36 |
| LR | 0.77 | 0.72 | 0.92 | 0.52 | 0.49 |
| NN | 0.72 | 0.62 | 1 | 0.24 | 0.41 |
| RF | 0.81 | 0.76 | 0.94 | 0.57 | 0.58 |
| XGB | 0.82 | 0.79 | 0.92 | 0.67 | 0.62 |
| **Older - Beat-AML2.0 and Leucegene** | | | | | |
| **Model** | **Accuracy** | **AUC** | **Sensitivity** | **Specificity** | **MCC** |
| DT | 0.67 | 0.6 | 0.32 | 0.88 | 0.25 |
| KNN | 0.62 | 0.5 | 0 | 1 | 0 |
| LR | 0.72 | 0.67 | 0.45 | 0.88 | 0.37 |
| NN | 0.69 | 0.65 | 0.48 | 0.82 | 0.32 |
| RF | 0.68 | 0.58 | 0.16 | 1 | 0.33 |
| XGB | 0.65 | 0.55 | 0.1 | 1 | 0.25 |
| **Older - Beat-AML2.0** | | | | | |
| **Model** | **Accuracy** | **AUC** | **Sensitivity** | **Specificity** | **MCC** |
| DT | 0.74 | 0.67 | 0.47 | 0.87 | 0.38 |
| KNN | 0.67 | 0.5 | 0 | 1 | 0 |
| LR | 0.74 | 0.66 | 0.42 | 0.9 | 0.37 |
| NN | 0.74 | 0.69 | 0.53 | 0.85 | 0.39 |
| RF | 0.76 | 0.63 | 0.26 | 1 | 0.44 |
| XGB | 0.72 | 0.58 | 0.16 | 1 | 0.33 |
| **Older - Leucegene** | | | | | |
| **Model** | **Accuracy** | **AUC** | **Sensitivity** | **Specificity** | **MCC** |
| DT | 0.48 | 0.5 | 0.08 | 0.91 | -0.01 |
| KNN | 0.48 | 0.5 | 0 | 1 | 0 |
| LR | 0.65 | 0.66 | 0.5 | 0.82 | 0.33 |
| NN | 0.57 | 0.57 | 0.42 | 0.73 | 0.15 |
| RF | 0.48 | 0.5 | 0 | 1 | 0 |
| XGB | 0.48 | 0.5 | 0 | 1 | 0 |

### Supplementary Table S6. Inversed models

| Younger kmers in older - Beat-AML2.0 and Leucegene | | | | | |
|---|---|---|---|---|---|
| **Model** | **Accuracy** | **AUC** | **Sensitivity** | **Specificity** | **MCC** |
| DT | 0.58 | 0.54 | 0.35 | 0.72 | 0.08 |
| KNN | 0.78 | 0.78 | 0.77 | 0.78 | 0.54 |
| LR | 0.8 | 0.79 | 0.74 | 0.84 | 0.58 |
| NN | 0.74 | 0.73 | 0.71 | 0.76 | 0.46 |
| RF | 0.81 | 0.83 | 0.87 | 0.78 | 0.63 |
| XGB | 0.79 | 0.76 | 0.65 | 0.88 | 0.55 |
| **Older kmers in younger - Beat-AML2.0 and Leucegene** | | | | | |
| **Model** | **Accuracy** | **AUC** | **Sensitivity** | **Specificity** | **MCC** |
| DT | 0.49 | 0.51 | 0.38 | 0.63 | 0.02 |
| KNN | 0.65 | 0.67 | 0.55 | 0.78 | 0.34 |
| LR | 0.7 | 0.71 | 0.66 | 0.76 | 0.42 |
| NN | 0.65 | 0.66 | 0.62 | 0.7 | 0.31 |
| RF | 0.41 | 0.45 | 0.23 | 0.67 | -0.11 |
| XGB | 0.63 | 0.67 | 0.46 | 0.87 | 0.35 |

**Supplementary Table S7. Summary of Cox model in younger patients**

```
Call:
coxph(formula = formule_y, data = data_y)

  n= 130, number of events= 44

              coef exp(coef) se(coef)      z Pr(>|z|)
age        0.03180   1.03231  0.01323  2.403 0.016252 *
TPM11      0.22863   1.25688  0.36932  0.619 0.535880
CLEC4OP1  -1.13703   0.32077  1.08071 -1.052 0.292749
TIAM11     0.11736   1.12453  0.41121  0.285 0.775330
GLCCI11    0.90886   2.48149  0.40907  2.222 0.026298 *
SLC29A21   1.47656   4.37785  0.38295  3.856 0.000115 ***
RACK11    -0.57063   0.56517  0.42401 -1.346 0.178366
NEIL11     0.14027   1.15059  0.41778  0.336 0.737054
LINGO31    0.66081   1.93635  0.36705  1.800 0.071809 .
IGKV2_241 -1.01137   0.36372  0.66922 -1.511 0.130722
---
Signif. codes:  0 ‘***’ 0.001 ‘**’ 0.01 ‘*’ 0.05 ‘.’ 0.1 ‘ ’ 1

          exp(coef) exp(-coef) lower .95 upper .95
age          1.0323     0.9687   1.00588     1.059
TPM11        1.2569     0.7956   0.60943     2.592
CLEC4OP1     0.3208     3.1175   0.03857     2.667
TIAM11       1.1245     0.8893   0.50228     2.518
GLCCI11      2.4815     0.4030   1.11305     5.532
SLC29A21     4.3779     0.2284   2.06679     9.273
RACK11       0.5652     1.7694   0.24619     1.297
NEIL11       1.1506     0.8691   0.50734     2.609
LINGO31      1.9364     0.5164   0.94309     3.976
IGKV2_241    0.3637     2.7494   0.09798     1.350

Concordance= 0.843  (se = 0.03 )
Likelihood ratio test= 69.23  on 10 df,   p=6e-11
Wald test            = 52.67  on 10 df,   p=9e-08
Score (logrank) test = 72.52  on 10 df,   p=1e-11
```

**Supplementary Table S8. Schoenfeld residuals for Cox model in younger patients**

| | chisq | df | p |
|---|---|---|---|
| age | 0.05863 | 1 | 0.809 |
| TPM1 | 0.95179 | 1 | 0.329 |
| CLEC4OP | 1.66884 | 1 | 0.196 |
| TIAM1 | 0.13693 | 1 | 0.711 |
| GLCCI1 | 1.11303 | 1 | 0.291 |
| SLC29A2 | 0.06483 | 1 | 0.799 |
| RACK1 | 0.14922 | 1 | 0.699 |
| NEIL1 | 0.34055 | 1 | 0.560 |
| LINGO3 | 3.93642 | 1 | 0.047 |
| IGKV2_24 | 0.00864 | 1 | 0.926 |
| GLOBAL | 8.51116 | 10 | 0.579 |

**Supplementary Table S9. Summary of Cox model in older patients**

```
Call:
coxph(formula = formule_o, data = data_o)

  n= 147, number of events= 98

       coef exp(coef) se(coef)     z Pr(>|z|)
age 0.06211   1.06408  0.01410 4.406 1.05e-05 ***
---
Signif. codes:  0 ‘***’ 0.001 ‘**’ 0.01 ‘*’ 0.05 ‘.’ 0.1 ‘ ’ 1

    exp(coef) exp(-coef) lower .95 upper .95
age     1.064     0.9398     1.035     1.094

Concordance= 0.621  (se = 0.033 )
Likelihood ratio test= 18.66  on 1 df,   p=2e-05
Wald test            = 19.41  on 1 df,   p=1e-05
Score (logrank) test = 20.03  on 1 df,   p=8e-06
```

**Supplementary Table S10. Schoenfeld residuals for Cox model in older patients**

| | chisq | df | p |
|---|---|---|---|
| age | 0.797 | 1 | 0.37 |
| GLOBAL | 0.797 | 1 | 0.37 |
